# A deterministic linear infection model to inform Risk-Cost-Benefit Analysis of activities during the SARS-CoV-2 pandemic

**DOI:** 10.1101/2020.08.23.20180349

**Authors:** John E. McCarthy, Bob A. Dumas, Myles T. McCarthy, Barry D. Dewitt

## Abstract

August 16, 2020

Risk-cost-benefit analysis requires the enumeration of decision alternatives, their associated outcomes, and the quantification of uncertainty. Public and private decision-making surrounding the COVID-19 pandemic must contend with uncertainty about the probability of infection during activities involving groups of people, in order to decide whether that activity is worth undertaking. We propose a deterministic linear model of SARS-CoV-2 infection probability that can produce estimates of relative risk for diverse activities, so long as those activities meet a list of assumptions, including that they do not last longer than one day. We show how the model can be used to inform decisions facing governments and industry, such as opening stadiums or flying on airplanes. We prove that the model is a good approximation of a more refined model in which we assume infections come from a series of independent risks. The linearity assumption makes interpreting and using the model straightforward, and we argue that it does so without significantly diminishing the reliability of the model.

## 1 Introduction

Coronavirus disease 2019 (COVID-19), caused by severe acute respiratory syndrome-coronavirus 2 (SARS-CoV-2), has caused a pandemic. As of August 2, 2020, the World Health Organization reported approximately 17.6 million cases and 680,000 deaths due to the disease [27]. Social distancing and shutting businesses have reduced the number of cases, but there is mounting pressure to reopen businesses. The purpose of this paper is to provide a model to estimate the relative infection risks of different activities. That information can allow decision-makers in industry and government to rank activities according to their relative risk of infection. In combination with an understanding of the benefits and costs of those activities, decision-makers can then make informed choices about whether, and if so, how to allow participation in previously forbidden activities.

Despite much ongoing research, there are many parameters of coronavirus disease that remain uncertain, such as the effective reproduction number of the virus given various characteristics of a population, or the precise effectiveness of various non-pharmaceutical interventions, or the significance of aerosol transmission [17, 7, 23]. Whereas much effort has been focused on determining these and other characteristics, many of which are needed to produce estimates of absolute risk of infection, such estimates are still uncertain. Nonetheless, policy decisions need to be made.

Risk-cost-benefit analysis provides one framework with which to analyze policy alternatives in order to inform policy decisions. In general terms, it aims to characterize the undesirable outcomes and the probabilities of those outcomes (i.e., the risks) for each decision alternative, the possibly uncertain costs of those alternatives, and their possibly uncertain benefits [11]. In its approach informed by behavioral decision research [11, 9, 13], the process involves not just normative analysis but also analysis to understand how the public perceives of the alternatives (i.e., descriptive analysis), and how to bridge the normative and descriptive perspective, when they differ (i.e., prescriptive analysis). Ultimately, the decision-maker also needs to perform a decision analysis with all of the information they have collected, which involves deciding on some decision rule to choose among the alternatives as characterized by their respective risks, costs, benefits, and the associated uncertainties.

In this study, we propose a deterministic model to estimate the relative risk of SARS-CoV-2 infection that we believe is useful for characterizing that risk for a large set of activities in both the private sector (e.g., attending a concert) and public sector (e.g., accessing government services in-person). That characterization also illuminates modifiable factors that can lower the risk of infection of a given activity. In combination with other information about the benefits and costs, the model provides a useful tool for anyone undertaking risk-cost-benefit analyses during the pandemic.

More specifically, we propose that when planning for activities that last no more than one day, we can use a model of infection probability that is linear in many potentially controllable variables, such as duration of the activity, density of participants, and infectiousness rate among the attendees. The advantages of a linear (deterministic) model are that it greatly simplifies analyses of different scenarios (for example, the effects of reducing density, or reducing the time spent in specific activities), and also allows comparison of relative risks across different events, even when the base parameters needed to estimate absolute risk are unknown.

This paper is organized as follows: in Section 2, we describe the assumptions of the model, and describe the model mathematically. In Section 3 we present an application, analyzing the risks of airplane travel. In Section 4 discuss how the model is useful, provide guidance for how it might be used, and address its limitations. Appendices provide mathematical details that we exclude from the main text.

## 2 The deterministic model

Here, we describe the deterministic linear model. We believe it is important to have a robust and well-formed, mechanistic, model of infection transmission, even if it contains many unknown parameters. As we will argue, this will allow us to draw inferences on *relative risks* even if we cannot quantify *absolute risks*. The availability of a model of infection transmission that is mechanistic will allow for comparative estimates of the consequences of specific policy choices. Confidence that a policy significantly reduces the risk of infection may be useful even in the absence of a reliable estimate of absolute risk.

We begin with some preliminary definitions, then describe the model’s assumptions, before describing the model proper.

### 2.0.1 Preliminary definitions

All the terms we define in this paper are included in Appendix 8. For now, we need the following terms:

By an *activity* we mean a well-defined set of interactions with clear bounds taking place over a period of time less than a day, for example a trip to a grocery store, or taking an airplane flight, or attending a sporting event as a spectator.

By the *participant* we mean a person attending the activity, whose probability of becoming infected we wish to model.

A *neighbor* at an activity is a person not in the participant’s immediate household who, for some part of the activity, is close enough to pose a risk of air-borne infection. We shall say the neighbor is in the participant’s *vicinity* if they are close enough to be a risk of infection. The precise nature of the vicinity is currently unknown; the CDC asserts that most infections are caused by individuals within 6 feet of each other [4], so a 6 foot radius may be an approximation for vicinity.

### 2.0.2 Assumptions

Like any model – whether deterministic or statistical – there are assumptions about the state of the world that are necessary for the model to apply. We state them here with some explanation, and discuss them in more detail in Section 4:

A1 Our first assumption is that the probability of infection is additive over sub-activities. This means that if one segments the activity into sub-activities, the probability of getting infected over the whole activity approximately equals the sum of the probabilities over each segment. Mathematically, this says that if we break an activity *A* up into *N* distinct sub-activities, *S*_1_*,…, S_N_* say, then the probability *p* of becoming infected during activity *A* satisfies

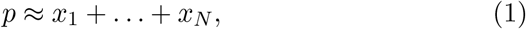

where *x_j_* is the probability of becoming infected during *S_j_*. We cannot actually expect exact equality in (1). Nonetheless – and this is an essential point – we can reasonably expect that the left-hand side and right-hand side of (1) agree with each other to within 10% or less. We give a mathematical proof of this assertion in Appendix 6.
A2 For each sub-activity *S_j_* the probability *x_j_* of infection is the sum over the forms of transmission of independent probabilities, each of which has a multiplicative form.
A3 If a neighbor is not infectious, there is 0 risk of infection from them.
A4 If a neighbor is infectious, the probability that they will infect the participant depends on the distance away, whether they are facing towards or away from the participant, mask usage, viral load in the neighbor, sneeze etiquette, air circulation, and other factors.
A5 The probability of infection from a neighbor is linear in the amount of time spent in their vicinity. See Appendix 6 for a justification of this assumption.
A6 The probability of infection in each segment is independent of the other segments.

Formally, the model does not need the following assumption, but it will be important when applying the model:

A7 There is no increased chance of infection from members of the participant’s immediate household engaging in the same activity, and we will ignore transmission from one’s immediate household members. (For example, sitting beside a household member at an activity will be treated as zero-risk).

### 2.1 Additivity over time

Assumption A1 is crucial to our study. Suppose we know an upper bound *π* on the chance of an individual becoming infected by SARS-CoV-2 over the course of a day’s activities. For some given activity *A*, such as attending a sporting event or taking an airplane flight, we break the activity up into temporally disjoint sub-activities, *S*_1_*,…, S_N_*. (For example: entering the stadium, walking to one’s seat, sitting and watching the event, going to a restroom, leaving the stadium). Suppose the probability of becoming infected in each subactivity *S_j_* is *x_j_*, and we wish to estimate *p*, the probability of becoming infected at some time during *A*. In Appendix 6 we prove that 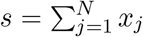 is a good approximation to *p*, and the smaller *π* is, the better the approximation. In particular, we show:

Theorem. *The following inequalities hold*:

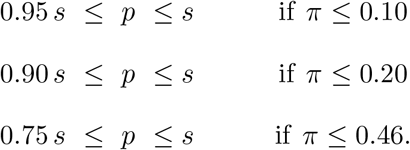

A simple example is useful to get some intuition about assumption A1 and the theorem. Suppose we were interested in the probability of rolling at least a single six when we roll three dice. That probability *p* is simple to calculate: it is 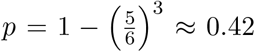, because the rolls are independent and the probability of rolling anything other than a six for a single dice is 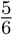. However, notice that, for this example, 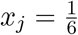 for all *j* ∈ {1,2,3}, and thus 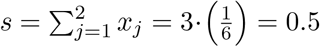. If we were not able to calculate *p* exactly, *s* is an approximation of *p* that is within 0.1 of the true probability. However, notice that, as the number of dice increases, *s* will quickly approach – and then equal, and then exceed – unity, despite a roll of six never being guaranteed.

A1 is crucial because, if we assume that

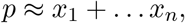

then it follows that:

- A given absolute reduction of risk in any segment *S_j_* has approximately the same overall impact on *p*.
- One can compare the relative risks from different activities, such as going grocery shopping, flying, or attending a sporting event, by analyzing sub-activities.

### 2.2 The full model

Putting together all of our assumptions, we wish to model the probability that a participant at an activity contracts SARS-CoV-2. Actual infection is understood to happen in one of three ways [28]:

- Airborne transmission from an infectious neighbor at the activity, either by droplets or aerosols.
- Touching a contaminated surface, and then touching the participant’s face before thoroughly washing the hands.
- Direct physical contact with an infectious person.

Each activity *A* is broken down into a sequence of segments *S_j_, j ∈ {*1*,…, N}*, disjoint sub-activities each of which can be thought of as a single uniform event, either as a single event (e.g. going to the restroom) or an event with constant parameters (e.g. sitting for some period of time with one neighbor 3 feet away, 2 neighbors 6 feet away, and no other neighbors within 10 feet).

Following A3, A4, and A5, for each segment *S_j_*, the probability that the participant becomes infected by air-borne transmission is the sum over every neighbor of [the probability the neighbor is infected] times [the probability the neighbor will cause the participant to be infected per unit time] times [the time spent in their vicinity]:

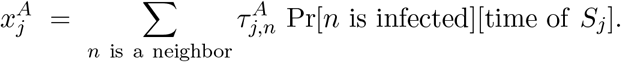

Here 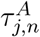 is the probability per unit time that given the configuration (distance away, orientation, mask-wearing or not, etc.) that if neighbor *n* is infected, they will infect the participant by air-borne transmission.

Similarly, the probability that the participant becomes infected by surface-born transmission from a surface they touch is [probability the surface is contaminated] times [probability they touch their face before washing their hands] times [probability that the touching leads to an infection]:

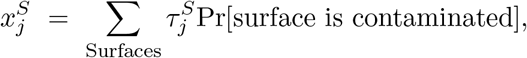

where 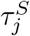 is the probability that the participant will convey the infection from the surface to themselves.

Finally, the probability that the participant becomes infected by direct contact with an infected neighbor is [probability neighbor is infected] times [probability of touching] times [probability of transmission]:

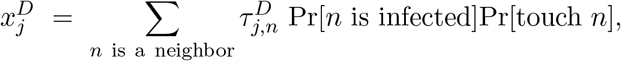

where 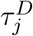 is the probability that if *n* is infected and the participant touches *n*, then infection will be transmitted.

Combining the above, we have (using A1, A2, and A6):

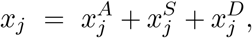

and

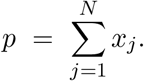

Our contention is that this model is strategically valuable even without knowledge of the parameters 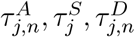.

## 3 Example: Air travel

Let us take travel on an airplane as an *activity*, as defined in Appendix 8. To use the model, we need to enumerate sub-activities *S_j_* that together make up the air travel activity, *A*. The sub-activities *S_j_* are:

1. boarding the plane
2. moving to and entering one’s seat
3. sitting on the plane for the duration of the flight
4. leaving one’s seat, and deboarding the plane.

The relevant parameters for this question, with sample values which can be changed, are:

1. *Position of seats in the plane*. We employ the seating arrangement used by United Airlines for the Boeing 737, which is available on United’s website [1]. Figure 1 shows the seating plan.
2. *Seating arrangement*. This can differ between scenarios, but for this example we will assume the plane is full.
3. *Time spent boarding, and traveling to one’s seat while on the plane*. We will assume the passenger stands in line for 10 minutes boarding, and takes 20 seconds to sit down at their seat once they reach the correct row on the plane. These values are estimates, and will vary according to airline boarding protocols.
4. *Order of seating*. We shall assume that the plane fills back to front, so that while walking to one’s seat, one does not pass already seated passengers.
5. *The distance apart people stand while boarding*. This can vary based on preventative measures taken by airlines; for this analysis, we will use 1.5 foot spacing.
6. *Duration of the flight*. This example will take flight duration as 180 minutes.
7. *Deboarding*, which will be modeled the same way as boarding for this example.
8. *How risk decays with distance*. There is much discrepancy in the literature as to this decay [7, 5]. Let us assume risk is inversely proportional to the square of distance from the source.

**Figure 1:**
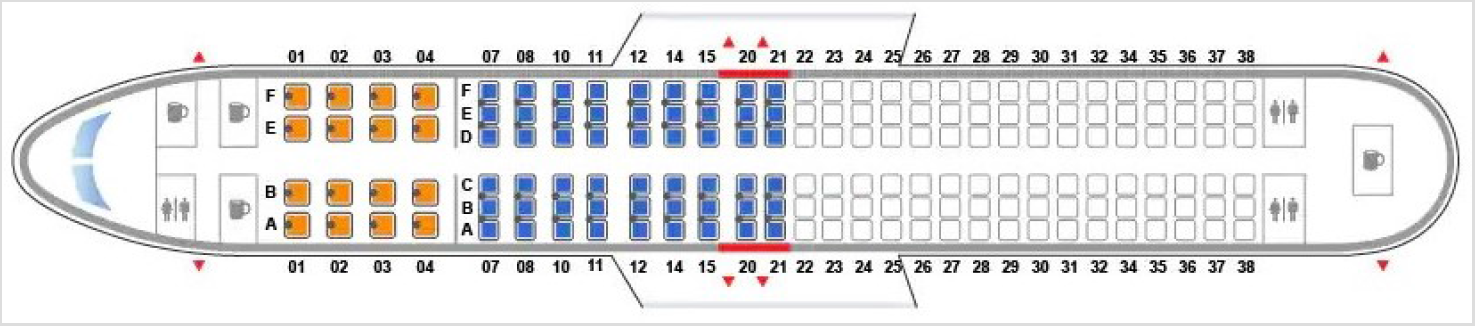
Boeing 737 Seating plan

We shall also assume for this example that there is no direct physical contact between participants and that all surfaces are disinfected.

For each sub-activity, a participant is exposed to some amount of risk from their neighbors. As we do not know absolute risks, we will quantify the risks of the various sub-activities using the hazard *×* exposure model described in the previous section. Given the analysis is one of *relative* risk, we do not have an absolute unit to use in the quantification of risk; thus, as a basic risk unit, we will use the risk of spending one minute at a distance of one foot from a stranger.

### 3.1 Boarding and deboarding

While boarding and deboarding, some number of strangers in the vicinity contribute to the risk a participant incurs. We assume that the boarding process arranges passengers linearly, and that the risk posed by strangers further than 6 feet away is negligible. The risk for boarding is obtained by summing the risk contribution of two strangers each 1.5 feet away, two strangers each 3 feet away, two strangers each 4.5 feet away, and two each 6 feet away for a duration of 10 minutes. Note that if parameter 5, separation distance, were to change, the number of srtangers for whom a risk contribution is calculated would also change.

Quantitatively, the risk is

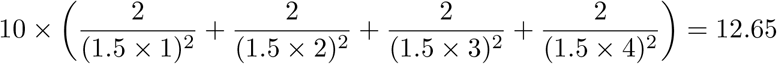

### 3.2 Entering and exiting seats

Entering and exiting seats and sitting on the plane are calculated similarly to each other, but rather differently from boarding and deboarding. While entering or exiting seats, on average, we calculate the risk contribution of each surrounding seat, sum them, and multiply by the duration taken to be 0.33 minutes. This works out to 1.14 for entering and for exiting. The cumulative risk for boarding, sitting, leaving the seat, and deboarding is thus 2 *×* (1.14 + 12.65) = 27.58.

### 3.3 Risk while seated

The risk calculation for sitting on the plane must account for the fact that some seats are spaced less densely on the plane than other seats. The methodology here is to calculate the average risk a participant incurs from their neighbors while seated. This value will not be the risk any individual passenger actually incurs, but is more accurate for the plane as a whole. The average risk value per minute is 1.84, so the average risk from sitting on plane for a three hour flight is 331.0. Thus, the average risk a participant incurs for this activity is 331.0 + 27.6 = 358.6.

### 3.4 Changing Parameters

Given the varying practices of the major airlines [16], the percentage of occupied seats is one parameter for which we are already seeing wide variation The results for similar scenarios are:

1. Airplane full, 1.5ft distancing while boarding Risk: 359
2. Middle seats empty, 3ft distancing while boarding Risk: 146
3. Airplane half full, 6ft distancing while boarding Risk: 100

Although the numbers 359, 146 and 100 are not in absolute units, they do show the relative effect of different possible mitigation strategies; other scenarios could be modeled similarly.

### 3.5 Other Sensitivity Analyses

In each of the above cases, we used the inverse square decay function to model the change in risk as distance to others changes. While the other parameters used in the model are measurable, the rate of decay of risk with distance has not been experimentally verified, and is quite uncertain. To see how sensitive to the decay function our conclusions are, we will try two very different decay functions of risk with distance. The first has a very slow exponential decay [7], and was based on averaging over many different studies; it concluded that each additional meter of distance decreased risk by a factor of 2.02. The second has a very rapid decay, based on simulations of droplet dispersion [5]. We shall refer to these as the Chu model and the Chen model, respectively. To normalize, we multiply the output of each function by a constant such that the risk at 3 feet is the same for each risk decay model. The risk values of each case with each other decay model are listed below.

**Table 1:** Relative risks with different decay assumptions.

|  | Case 1 | Case 2 | Case 3 |
| --- | --- | --- | --- |
| Inverse Square | 359 | 146 | 100 |
| Chu Model | 405 | 219 | 159 |
| Chen Model | 1231 | 181 | 121 |

There are many similarities across the different decay rates. In all cases, the vast majority of risk is incurred by seating, and Case 3 is about 2*/*3 the risk of Case 2. The very large discrepancy in Case 1 is explained by the extreme sensitivity of the Chen model at close ranges. According to this model, moving from 0.2 meters to 0.3 meters reduces risk by a factor of 88. If a sensitive model such as the one presented by Chen et al. is most accurate, it will be important to be aware of the interval or intervals where risk drops rapidly.

## 4 Discussion

In this paper, we have described a deterministic linear model for estimating the relative risk of infection by SARS-CoV-2 during an activity that lasts less than a day. Even without being able to estimate absolute risks, it allows decision-makers to rank activities according to how much risk of infection they pose to the public; in combination with knowledge about those activities’ costs and benefits, decision-makers can make more informed choices about whether, and how, to allow people to participate in currently forbidden activities [14]. Crucially, as demonstrated in the example, an analysis using this model also reveals which segments of the activity pose the greatest risk. When these are modifiable, stakeholders can act to lower the risk.

In contexts where the model applies, it has significant policy value, despite only being able to calculate relative risks:

- The model is linear in time. Therefore, engaging in an activity for twice as long doubles the chance that a participant becomes infected. Boarding airplanes, for example, can be done in much more efficient ways than is currently the norm [10]. Optimizing the boarding process so that passengers spend less time close to neighbors will reduce their infection risks.
- The model is linear in the proportion of attendees at an activity who are infectious. This number in turn is the product of two numbers: the proportion of the population who are infectious (which will vary over time) and the probability that an infectious person will not self-isolate. The latter number can be reduced by public health education, by testing and contact tracing, and by health checks.
- The model is linear in the probability that a given neighbor will cause an infection. In [22] the authors find that home-made masks block 95% of air-borne viruses, medical masks block 97%, and N95 masks block 99.98%. That study was a mechanical simulation using nebulizers and did not distinguish in application between a potentially infectious person wearing a mask to reduce their probability of transmission, and an uninfected person wearing a mask to reduce their probability of infection. In [6], the effectiveness of masks in practice is considered. It is reasonable to assume that everybody wearing masks reduces airborne transmission by some factor. In [22] they do a meta-analysis, and find that wearing masks reduced risk, with high uncertainty in the amount, but their point estimate was a reduction to 23% of the non-mask wearing risk when using non-respirator masks (and a reduction to 4% using respirators).
- The model is linear in density of neighbors. Reducing the number of neighbors at a given distance by 50% reduces by 50% the chance of air-borne infection. Leaving seats open, and clustering only members of the same household, will reduce the risk of air-borne transmission.
- For surface-borne infections, the model is linear in the probability that the surface is contaminated. Doubling the cleaning frequency will approximately halve the probability that the surface is contaminated.

These linearities allow for comparisons among different scenarios, and comparisons across different activities.

Of course, deciding whether and how to relax restrictions on activities requires understanding the public’s perceptions of the risks, costs, and benefits of doing so. Formally equivalent risks could be perceived differently, in ways that might seem irrelevant to a risk analyst but would impact the decisions of potential participants in an activity [15, 25]. Furthermore, so long as there is some nontrivial amount of virus in the community, activities involving large numbers of people will almost certainly lead to some eventual transmission. Decision-makers need to evaluate the testing and contact-tracing infrastructure of the jurisdictions where the activities are located to determine whether that transmission can be contained, given that any transmission due to the activity is a burden not only to the participants, but also to the entire community. Decision-makers need to invest in empirically-tested risk communication so that participants understand the risks accurately and the public at large understands why that risk is judged to be acceptable by policymakers. There are already detailed frameworks for engaging the public about scientific and technical risks and undertaking an analytical-deliberative process to develop a plan that is widely endorsed among stakeholders [26]. There is no reason these frameworks cannot be adapted for the SARS-CoV-2 pandemic [12].

The model should be conceived as a tool in a bottom-up-analysis – there is no one-size-fits-all approach to the problem. Each activity has specific characteristics only known to those involved with the activity; even enumerating them can require specialist knowledge. The June 2020 report describing how the entertainment industry can safely return to work [8], jointly authored by the major unions of that industry, is an example of that synthesis of modeling, industry knowledge, and risk communication.

Using the example above of the airplane analysis, one can see the relative benefits of different mitigation strategies. Making masks mandatory, and enforcing this rule, is clearly the most cost-effective strategy. Keeping the middle seat vacant unless there is a party of three travelling together at least halves the risk, under a very wide range of decay assumptions. Managing boarding is less costly, but the total impact will be lower since it takes up a small part of the total flight time.

### 4.1 Limitations

Like any model, the model described here depends on its assumptions. In our view, the most problematic assumptions are A2 and A6, which require independence. That could fail, if, for example, infection requires a certain minimum threshold of exposure. Similarly, A7, the assumption that those in one’s household pose no threat, could be problematic. It is probably true during the activity; however, the presence of family members could increase the risk of exposure after the activity when one returns home, given, e.g., their separate trips to the washroom or through the turnstile during the activity. The importance of these extra exposures is an empirical question, and a function of the relative risk of those sub-activities done individually, and protective actions taken after the event (e.g., social distancing, hand-hygiene, proactive testing etc.).

More importantly, the model assumes that the risk of infection is a function of the background risk in the population of the activity’s jurisdiction. However, that assumes the subpopulation of potential participants does not have more virus prevalence than the community at large. Whether that is true is also an empirical question. For example, are those who would choose to attend a stadium concert during a pandemic more or less likely to participate in protective actions that lower their overall risk of virus infection or of virus transmission? Part of the empirical study of the public’s risk perceptions would need to include an appraisal of that question.

Furthermore, the model has a specific definition of “activity,” and it is crucial that the definition is clear to those who would use the model. For instance, considering a semester on a university campus as *∼* 90 separate one-day activities would not be an appropriate use of the model, because the population on campus from one day to the next is almost identical. Unless participation in an activity incurs no additional risk for a participant beyond their usual activities, decision-makers would need to consider how to limit individual participation; for example, in limiting the number of sports games one can attend in a given time period. Otherwise, the risk of transmission among participants will surely increase over the average risk in the community as previously-shuttered activities become a part of their day-to-day life but not of the lives of their fellow community members. The feasibility of such controls should be considered during the decision-making process.

For analytical purposes, our example shows how sensitive many analyses will be to uncertainty about how the probability of infection decays with distance, including the threat of long-range exposure [7, 5, 3]. As long as estimates for those values vary widely in the literature, decision-makers may need to be conservative in cases where an analysis is highly sensitive to changes in the relevant parameters.

## 5 Conclusion

The SARS-CoV-2 pandemic has restricted the activities of every person in the world. As governments and businesses try to decide how to reopen society, they need an analytical framework with which to make reasoned decisions. While much of the modeling done to date has focused on estimating the parameters needed to calculate the absolute risk of SARS-CoV-2 infection, here, we focus on estimates of relative risk. Such a model should allow decision-makers to rank the risk of activities. Combined with an accounting of their benefits and costs, decision-makers would have the information necessary to make informed decisions.

## Data Availability

No external data

## 6 Appendix: Additivity in time

### 6.1 Mathematical derivation of approximate additivity

Let us assume that an activity *A* is decomposed into *N* segments, called *S*_1_*,…, S_N_*, and each segment *S_j_* has some risk *x_j_* of causing infection. We further assume that these risks are statistically independent of each other. Then the probability *p* of being infected at some time during *A* is

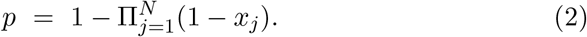

Let

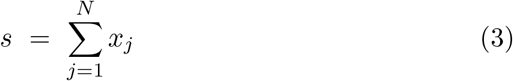

Then we claim that

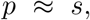

where the symbol *≈* means “is approximately equal to”. Indeed we claim that 1 *− e^−s^s*

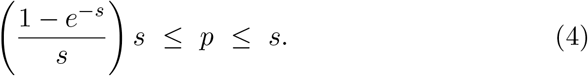

To see (4), we use the arithmetic-geometric inequality to show

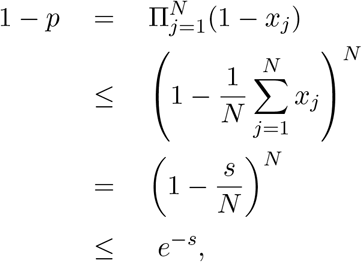

which yields the left-hand inequality in (4).

The right-hand inequality follows from

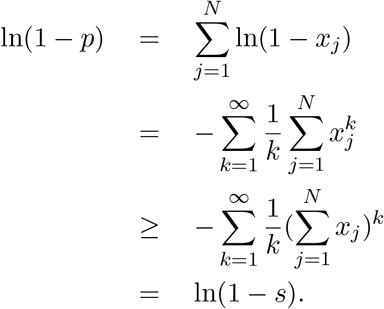

The correction factor between *s* and *p* is 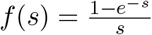. This is a decreasing function of *s*, which has a right-hand limit of 1 as *s* tends to 0. Since *s ≤ −* ln(1 *− p*), we have

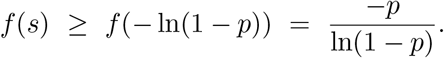

So our conclusion is that we always have

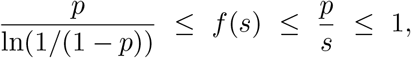

justifying the claim that *p ≈ s*. For representative values of *f*(*s*) and *g*(*p*) = *−p/* ln(1 *− p*), see Tables 2 and 3.

**Table 2:** 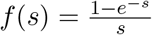 as a function of *s*

|  |  |  |  |  |  |  |  |  |
| --- | --- | --- | --- | --- | --- | --- | --- | --- |
| $s$ | 0 | 0.05 | 0.1 | 0.15 | 0.2 | 0.3 | 0.4 | 0.5 |
| $f(s)$ | 1.00 | 0.98 | 0.95 | 0.93 | 0.91 | 0.86 | 0.82 | 0.79 |

**Table 3:** 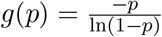 as a function of *p*

|  |  |  |  |  |  |  |  |  |
| --- | --- | --- | --- | --- | --- | --- | --- | --- |
| $p$ | 0 | 0.05 | 0.1 | 0.15 | 0.2 | 0.3 | 0.4 | 0.5 |
| $g(p)$ | 1.00 | 0.97 | 0.95 | 0.92 | 0.90 | 0.84 | 0.78 | 0.72 |

### 6.2 Estimating an upper bound on *p*

As we saw in Subsection 6.1, how good the approximation *p ≈ s* is depends on how close *f*(*s*) is to 1. How can we measure this?

We use the assumption that the activities we are considering last less than a day. While some activities are more risky than others, we further assume that all the events will be designed so that the total risk of an infected individual spreading the infection is no greater than it was at the beginning of the pandemic before any social distancing measures were in place. So we get an upper bound

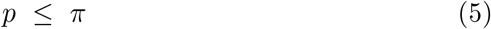

where *π* is the probability that before social distancing, an infectious individual would infect a susceptible individual over the course of one day. Note that for many activities, it is reasonable to assume that *p* is much less than *π*, thus tightening the estimation in Subsection 6.1.

Since an infected individual can infect multiple susceptibles, the expected number they would infect over the course of the day would be slightly higher, namely 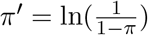 if one assumes a Poisson distribution. (This is a small adjustment that will not materially affect our conclusion, so the reader can ignore it.)

Using a standard SIR model at the early stage of an infection, the proportion of the population that is susceptible is close to 1. So the proportion of the population that is infected will grow exponentially, like *e^κt^* for some rate *κ*, where 1+*π*′ = *e^κ^*. The doubling time *τ* is the time at which *e^κτ^* = 2, so *κ* = ln(2)*/τ*. Thus we get

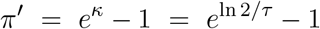

and

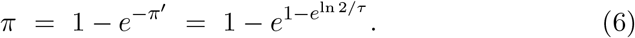

What is *τ*? In [24] they estimate that the doubling times in Chinese provinces in the period January 20 - February 9 2020 ranged from 1.4 days (95% CI 1.2-2.0) in Hunan province to 3.1 days (95% CI 2.1-4.8) in Xinjiang province.

In [21], the authors estimate the doubling time in Italy in March 2020 to be 3.4, 5.1 and 9.6 days in the first, second and third ten day periods of the month.

In [19], the authors estimate the probability of infection in a crowded zone (summed over all neighbors in the vicinity) to be 1.8% per hour, and to be 0.18% and 0.018% in moderate and uncrowded zones, using data from the cruise ship Diamond Princess. If we assume at most 12 hours spent in crowded zones per day on the cruise ship, this would yield the estimate

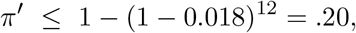

**Table 4:** *π* as a function of *τ*, the doubling time.

| $\tau$ | $\pi$ | $f(\pi)$ | $g(\pi)$ |
| --- | --- | --- | --- |
| 1 | 0.63 | 0.74 | 0.63 |
| 1.4 | 0.47 | 0.80 | 0.74 |
| 2 | 0.34 | 0.85 | 0.82 |
| 3 | 0.23 | 0.89 | 0.88 |
| 4 | 0.17 | 0.92 | 0.91 |
| 5 | 0.14 | 0.93 | 0.93 |
| 6 | 0.12 | 0.94 | 0.94 |
| 7 | 0.10 | 0.95 | 0.95 |
| 8 | 0.09 | 0.96 | 0.96 |

 which in turn from (6) gives

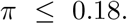

In [2], the authors use an SEIR model on the data from [20], and take into account the incubation period (6 days) recovery period (14 days) and a mortality rate of 1%. They assume that transmission rates are the same in asymptomatic and symptomatic states, and get a value of *π* that is 0.126. This follows from their equation

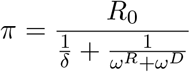

and using their values *R*_0_ = 2.5, *ω^R^* = 1*/*14 is recovery rate from symptomatic to recovered, and *ω^D^* = .01 is the mortality rate.

The value *R*_0_, the number of new people infected per infectious person, is widely reported by time and geographic region – see e.g. [18] for estimates of *R*_0_ by U.S. state. If one makes the more conservative estimate that only asymptomatic carriers will be circulating, and using the same 6 day incubation period, then one gets the bound

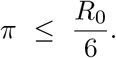

With an estimate of 2.22 for pre-mitigation *R*_0_ in the U.S. [18], this gives the bound

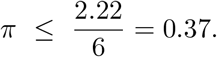

## 7 Appendix: Refinement to additivity over segments

One can refine the analysis in Appendix 6. Let us assume (2) and (3) both hold, and that we have segmented the activity into sufficiently small pieces that each *x_j_ ≤ ε* for some small *ε* that we assume satisfies 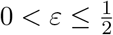.

Then we can tighten the bounds in (4) to

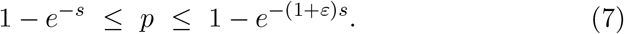

We omit the proof.

## 8 Appendix: Definitions

By the *participant* we mean a currently non-infected person attending the activity, whose probability of becoming infected we wish to model.

A *neighbor* at an activity is a person not in the participant’s immediate household who, for some part of the activity, is close enough to pose a risk of air-borne infection. We shall say the participant is in the neighbor’s *vicinity* if they are close enough to become infected.

*π* is the probability that withour social distancing, an infectious individual would infect at least one susceptible individual over the course of one day.

*π*′ is the expected number of new infections per day caused by an infectious individual without social distancing.

*τ* is the doubling time of the infection.

## 9 Conflict of Interest Statement

J.E.M., B.A.D. and M.T.M. received funding as consultants for Delaware North, a company that may be affected by the research reported in the paper.

